# Virtual Care and Medical Cannabis Access: A Geospatial Study of Telemedicine’s Role in Reducing Socioeconomic Disparities

**DOI:** 10.1101/2024.12.05.24318514

**Authors:** Mitchell L. Doucette, Emily Fisher, Dipak Hemraj, Mark Kasabuski, Junella Chin

**Affiliations:** Health Economics and Outcomes Research Division, Leafwell, Miami, FL

## Abstract

**Introduction:** Telemedicine has the potential to improve healthcare access and reduce disparities. We examined whether the incidence rate of medical cannabis patients (MC) was associated with concentrated disadvantage in Pennsylvania in 2022, accounting for a population of patients approved through telemedicine.

**Methods:** This zip code-level analysis examined associations between the Concentrated Disadvantage Index (CDI) and two outcomes: (1) the number of telemedicine-approved MC patients, as obtained from a specific telemedicine provider, and (2) the number of all other MC patients, calculated by subtracting the number of telemedicine-approved patients from the total number of MC patients at the zip code-level. Total counts of MC patients and in-office MC providers for Pennsylvania were sourced from the Pennsylvania Department of Health, while CDI data were derived from the 2022 American Community Survey. We used multivariate negative binomial regression models with population offsets and robust standard errors, adjusting for spatial autocorrelation through spatial lag adjustments.

**Results:** The CDI was not associated with the incidence rate of telemedicine-approved MC patients (IRR = 0.962; *p* = 0.355), but it was significantly negatively associated with the incidence rate of all other MC patients (IRR = 0.904; *p* = 0.000). The density of in-office MC providers was significantly associated with the incidence rate of all other MC patients but not with telemedicine-approved patients. Spatial factors, including autocorrelation, significantly influenced the distribution of both groups of patients.

**Discussion:** These findings suggest that telemedicine plays a crucial role in improving access to MC for socioeconomically disadvantaged areas. The lack of a significant association between the CDI and telemedicine-approved MC patients highlights the ability of telemedicine to bypass barriers such as provider scarcity and transportation challenges. By facilitating remote consultations and approvals, telemedicine ensures access for patients who might otherwise face difficulties obtaining MC.

## 1 Introduction

### 1.1 Medical Cannabis in the United States: Policies and Promises Unkept

In the United States, 38 states and the District of Columbia have legalized cannabis for medical use (National Conference of State Legislatures, 2024). All states where cannabis is medically available have developed state-administered medical cannabis (MC) programs. State regulatory frameworks for MC vary, influencing which conditions qualify for treatment and the demographic profiles of users (Boehnke et al., 2024; Fairman, 2016). Prospective patients must obtain certification from a qualified physician, verifying their diagnosis of one of these conditions to become eligible for MC use. As of 2022, there were nearly 4.1 million registered MC patients in the U.S., a 33% increase from the previous year (Boehnke et al., 2024).

Current research suggests that MC has beneficial properties (National Academies of Sciences, 2017). While it is not without risks, MC has been shown to assist in the discontinuation of high-risk medications (Bradford and Bradford, 2016, 2017, 2018; Bruce et al., 2021b; Charoenporn et al., 2023; O’Connell et al., 2019; Purcell et al., 2019). It has been shown to increase sleep quality and quality-of-life for people suffering from various conditions including chronic pain, post-traumatic stress disorder, and epilepsy (AminiLari et al., 2022; Bonn-Miller et al., 2022; Bruce et al., 2021a; Cahill et al., 2021; Chan et al., 2017; Drost et al., 2017; Erridge et al., 2022, 2023; Harris et al., 2022; Mangoo et al., 2022; Nicholas et al., 2023; Pillai et al., 2022). Examining the impact of medical and recreational cannabis laws through a societal lens shows evidence that MC laws do not generate negative economic outcomes or lead to increased criminal activity (French et al., 2022; Ghimire, 2020; Ullman, 2017). However, the increased access to MC resulting from policy reform, and thus its potential benefits, have not been realized equally across racial and ethnic groups (American Civil Liberties Union, 2020; Martins et al., 2010).

Despite the recent push towards making cannabis legal for medical and recreational purposes within the U.S., the harms surrounding cannabis criminalization continue to disproportionately impact Black and Hispanic people. A report by the American Civil Liberties Union (2020) notes that the war on cannabis continues despite law changes. This report, and others specific to New York, find persistent racial disparities in cannabis possession arrests in states that legalized cannabis (Drug Policy Alliance, 2021).

These disparities exist despite policymakers making concerted efforts to pass cannabis reform legislation that includes diversity requirements in MC and adult-use laws. These measures aim to address social equity by ensuring that communities disproportionately affected by previous cannabis prohibition have opportunities to participate in the legal cannabis market (New Jersey Cannabis Regulatory Commission, 2024; State of Connecticut, 2024). But the efforts to implement diversity requirements in MC and adult-use cannabis laws have shown mixed progress as the legislative process has been slow, with many bills still awaiting committee approval (WHYY, 2024). The pace of legislative action and the complexity of regulatory frameworks continue to challenge the full realization of these diversity goals.

### 1.2 Telemedicine in the United States

Telehealth has been recognized for its potential to deliver efficient and cost-effective care by reducing healthcare costs through decreased medication misuse, unnecessary emergency department visits, and prolonged hospitalizations (Ebbert et al., 2023; Gajarawala and Pelkowski, 2021; Shaver, 2022). Patient preferences indicate a high likelihood of using telemedicine for medication refills, preparing for visits, reviewing test results, and receiving education, showcasing its effectiveness in various healthcare services (Ebbert et al., 2023). Furthermore, telemedicine interventions have shown promise in managing chronic conditions, suggesting potential long-term benefits for patient outcomes and healthcare cost reduction (Omboni et al., 2020; Tchero et al., 2019).

Telehealth also plays a crucial role in increasing access and alleviating disparities in healthcare. It offers significant benefits for socially or economically disadvantaged populations, such as those in rural areas, who face greater barriers to accessing traditional in-person care (Ebbert et al., 2023; Mahtta et al., 2021). However, disparities in telemedicine utilization persist, particularly among racial and ethnic minorities, individuals with low socioeconomic status, and those with limited technological access (Mahtta et al., 2021; Marcondes et al., 2024; Shaver, 2022).

### 1.3 Medical Cannabis Access and Telemedicine

To date, there is a dearth of research evaluating whether access to medical cannabis has been equitably distributed. One study, Cunningham et al. (2022), examined the geographic distribution of in-office MC certifying providers in New York. The study found that for every 10% increase in the percent of Black residents, neighborhoods were 5% less likely to have at least one in-office MC provider. Conversely, they found that for every 10% increase in the percent of residents with a bachelor’s degree or more, neighborhoods were 30% more likely to have at least one in-office MC provider. These findings suggest that in-office qualified providers are unevenly distributed across socioeconomic groups, with significant implications for accessing MC (Cunningham et al., 2022).

Telemedicine offers a potential solution to address the uneven geographic distribution of MC providers. Its use has recently increased in popularity (DeWitt, 2020; Pankratz, 2023). Currently, only two states, Rhode Island and Utah, do not allow for telemedicine consultations for qualifying a patient for MC. Although limited research compares the two consultation modes, one study, a doctoral thesis, found that telemedicine visits were as effective as in-office visits for reducing pain among chronic pain patients treated in a MC clinic during the COVID-19 pandemic, highlighting the potential for telemedicine as an alternative mode of care (Pankratz, 2023).

### 1.4 Current Contribution

We sought to understand whether telemedicine consultation services addressed existing disparities in MC access found in the Cunningham et al. (2022) study of New York. To do so, we explored the relationship between telemedicine-approved MC patients and concentrated disadvantage across Pennsylvania zip codes in 2022, accounting for spatial distribution and in-office MC providers. Pennsylvania legalized MC on April 17, 2016. The first licensed sales occurred on February 15, 2018. As of July, 2023 the Pennsylvania Department of Health (PA-DoH) announced a total of 942,231 registered patients and caregivers.

## 2 Methods

This cross-sectional study used publicly available 2022 data from PA-DoH, proprietary Leafwell data, and geospatial data from the National Historical Geographic Information System (NHGIS). The analysis was conducted at the zip code-level, examining the associations between concentrated disadvantage and counts of patients, controlling for the density of in-office certifying medical providers.

### 2.1 Study Variables

We examined two primary outcomes: counts of telemedicine-approved MC patients from Leafwell and counts of all other MC patients by zip code (Leafwell, 2024). We identified telemedicine-approved patients through the Leafwell Patient Database (LPD). Leafwell operates in 36 states and advertises on internet search engines and digital media to connect potential MC patients with physicians in their state. After a physician deems a patient qualified for MC, Leafwell assists patients in obtaining their medical card. Leafwell patients are asked to fill out a baseline questionnaire, providing their zip code of primary residence. Data from the LPD have been used recently to describe common primary qualifying conditions for medical cannabis (Doucette et al., 2024b,a) as well as estimate the impact of medical cannabis treatment on healthcare utilization (Doucette et al., 2024c)

To achieve our research aims, we accessed and analyzed LPD data from January 1, 2022, to December 31, 2022. We identified all patients approved in Pennsylvania in 2022 and obtained their zip code information. For patient confidentiality, only de-identified data from the LPD was shared with internal researchers. Researchers did not have access to identifiable data. This project received exempt status from an external, third-party IRB, BRANY (IRB Number: IRB00000080). Patients consented to the use of their questionnaire data in aggregate form as part of the Leafwell terms of service.

Data on all other MC patients in Pennsylvania were obtained through a release resulting from the court case Department of Health v. Spotlight PA, Commonwealth Court of Pennsylvania, No. 660 C.D. 2021 (Mahon, 2023a). Spotlight sued the PA-DoH for de-identified data related to MC patients. As a result, the PA-DoH provided anonymized MC patient data for 2017 through 2022 to Spotlight, which subsequently published the data. We downloaded the Spotlight data for 2022, which provided the zip code of patients (Mahon, 2023b,a). We aggregated both sets of patient data (LPD and PA-DoH) to the zip code-level. We then subtracted the counts of telemedicine-approved patients from the counts of total Pennsylvania patients to create the outcome variable all other MC patients.

Our primary independent variable was the concentrated disadvantage index (CDI). GIS zip code boundaries, as well as zip code-specific demographic data necessary to compute the CDI, were downloaded from the Integrated Public Use Microdata Series’ (IPUMS) NHGIS (Manson et al., 2023). We downloaded data for the year 2022. The IPUMS-NHGIS provides summary tables of the 2022 American Community Survey (5-year average 2018-2022). We did not use data from 1-year or 3-year estimates, as these are not available at the zip code-level.

We defined the CDI variable following established literature (Jing et al., 2022). Items were combined into an index by taking the average of their z-scores, per established literature. A higher value of the CDI variable indicates more concentrated disadvantage within a zip code. We also included in the analysis a covariate representing the percentage of the population that was white non-Hispanic. White non-Hispanics have been shown to utilize MC at higher rates compared to other race/ethnicities (Fairman, 2016; Mahabir et al., 2020).

We included counts of in-office MC providers as an additional covariate in our models. We obtained counts of in-office MC providers from the State of Pennsylvania’s MC program. This information is publicly available through the PA-DoH (Pennsylvania Department of Health, 2024). We downloaded the information, cleaned, and aggregated the counts of in-office MC providers to the zip code-level for the analysis.

### 2.2 Statistical Approach

The unit of analysis was the zip code-level. To examine whether concentrated disadvantage was different for telemedicine-approved patients versus all other patients, we first conducted a multi-variate negative binomial regression including all covariates in the model. We also conducted an analysis to estimate whether spatial autocorrelation impacted our findings. To achieve this, we first defined the spatial relationships between the observations, creating a spatial weight matrix using the four nearest neighbors. We then conducted Moran’s I tests for our two primary outcomes to test for autocorrelation. We extracted the residuals from the initial regression model, calculated a spatial lag, and fit them into the spatial regression model, which included the covariates mentioned above and lagged residuals to account for spatial dependence. For both the initial model and the spatial regression model, we included a zip code-specific population offset to express our estimates as incidence rate ratios (IRR). Both models also included standard errors clustered at the zip code-level to adjust for potential correlation within clusters. All analyses were conducted using R version 4.3.1 (R Core Team, 2024). The multivariate negative binomial regression models were conducted using the glm.nb function in the R package MASS (Venables and Ripley, 2002).

## 3 Results

Of the 2,167 total zip codes associated with Pennsylvania, we identified 1,458 standard zip codes, excluding all PO boxes and unique zip codes. Of the 1,458 zip codes, 53 had a population of zero. Therefore, we excluded these zip codes for a total of 1,405 remaining in the analyses.

We provided figures examining the distribution of telemedicine-approved MC patients, all other approved MC patients, in-clinic MC providers, and the CDI, respectively in Figures 1, 2, 3, and 4. Each figure provides zip code-level information for Pennsylvania, specifically highlighting Philadelphia and Pittsburgh. Examining Figures 1 and 2, we see that there is some overlap in telemedicine-approved and all other patients, but with a greater distribution for all other patients.

**Figure 1:**
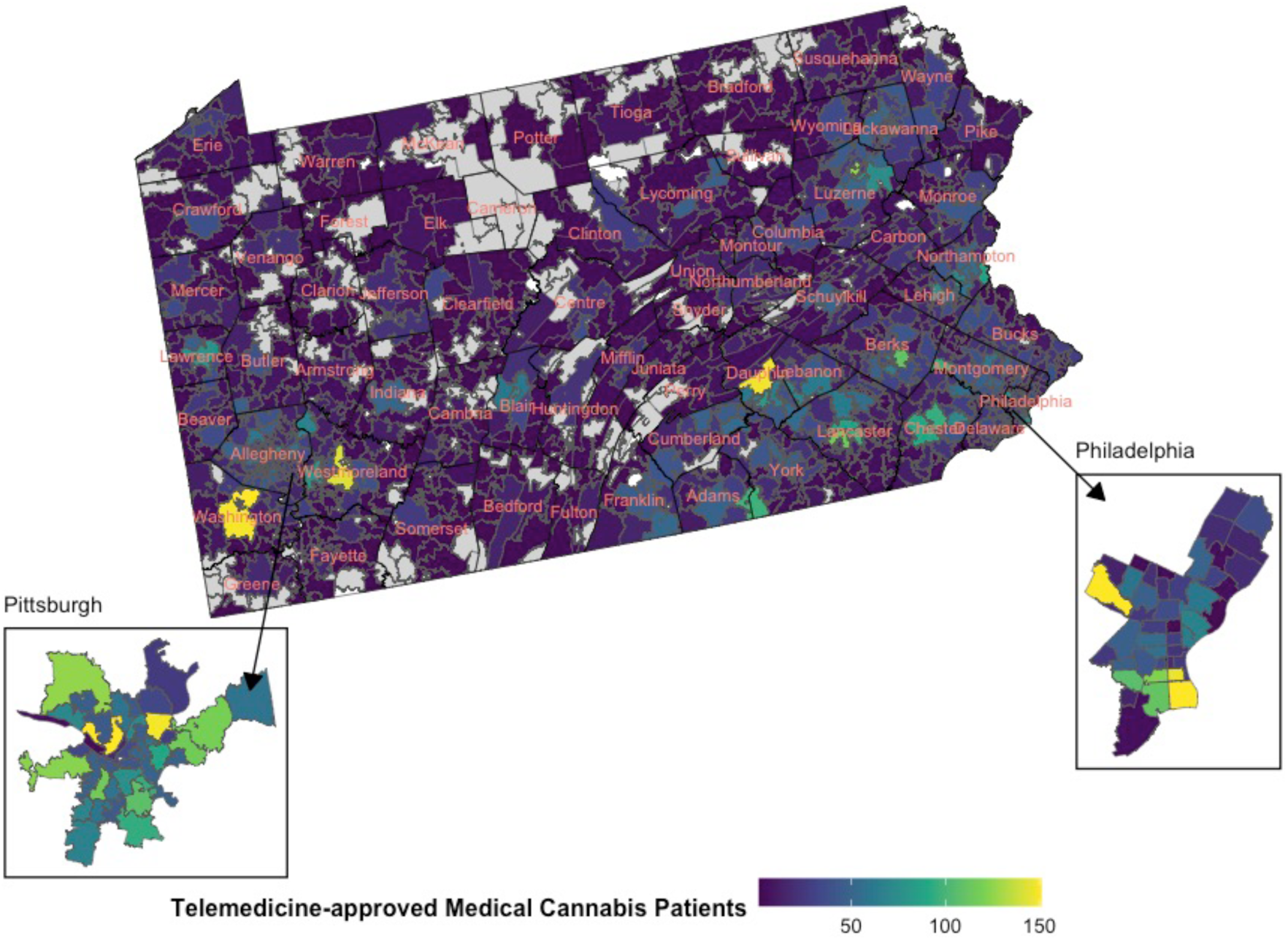
Distribution of telemedicine-approved patients within Pennsylvania in 2022. Light gray indicates no patients were present in the zip code. White indicates limited data on population.

**Figure 2:**
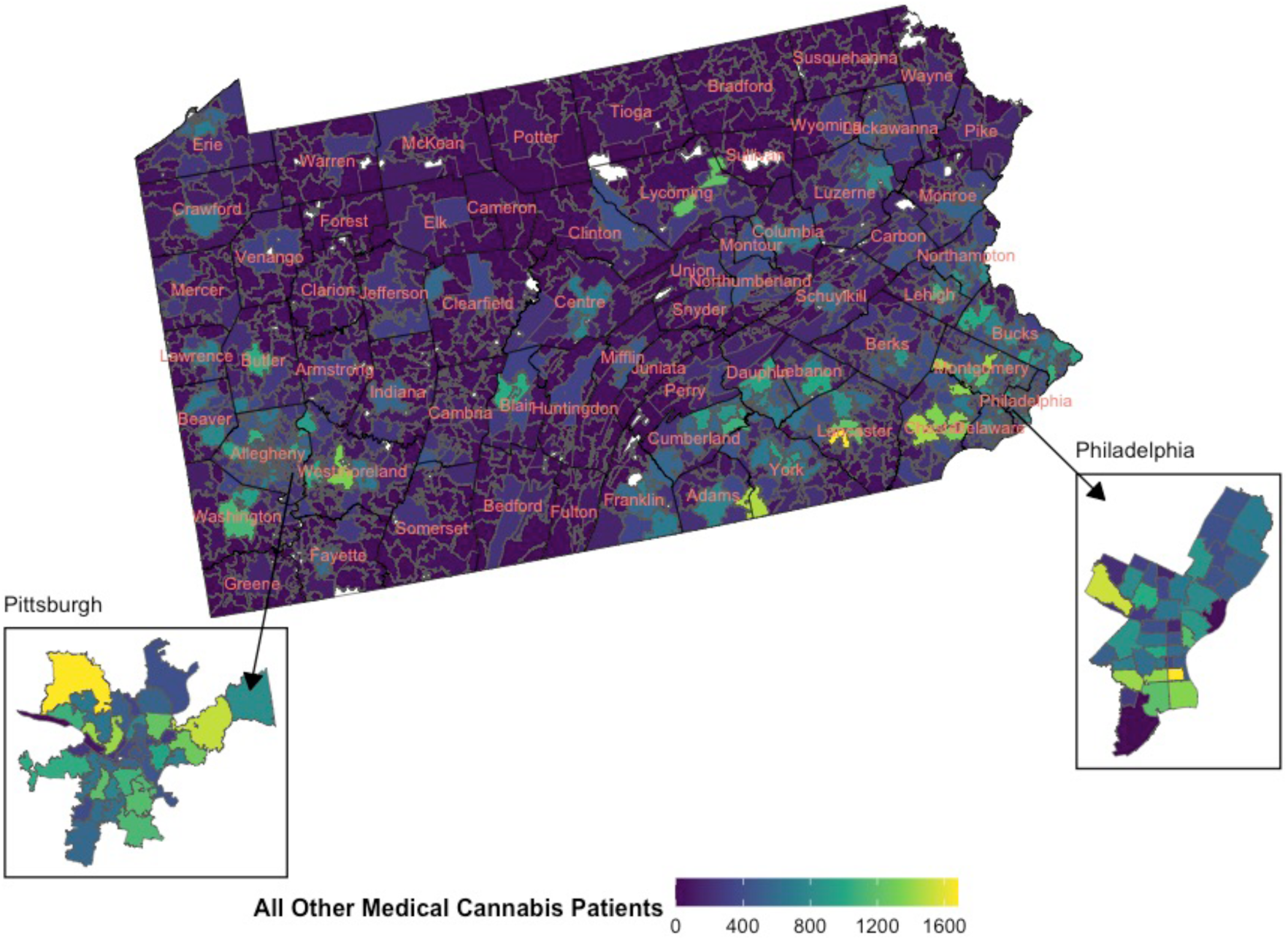
Distribution of all other patients within Pennsylvania in 2022.Light gray indicates no patients were present in the zip code. White indicates limited data on population.

**Figure 3:**
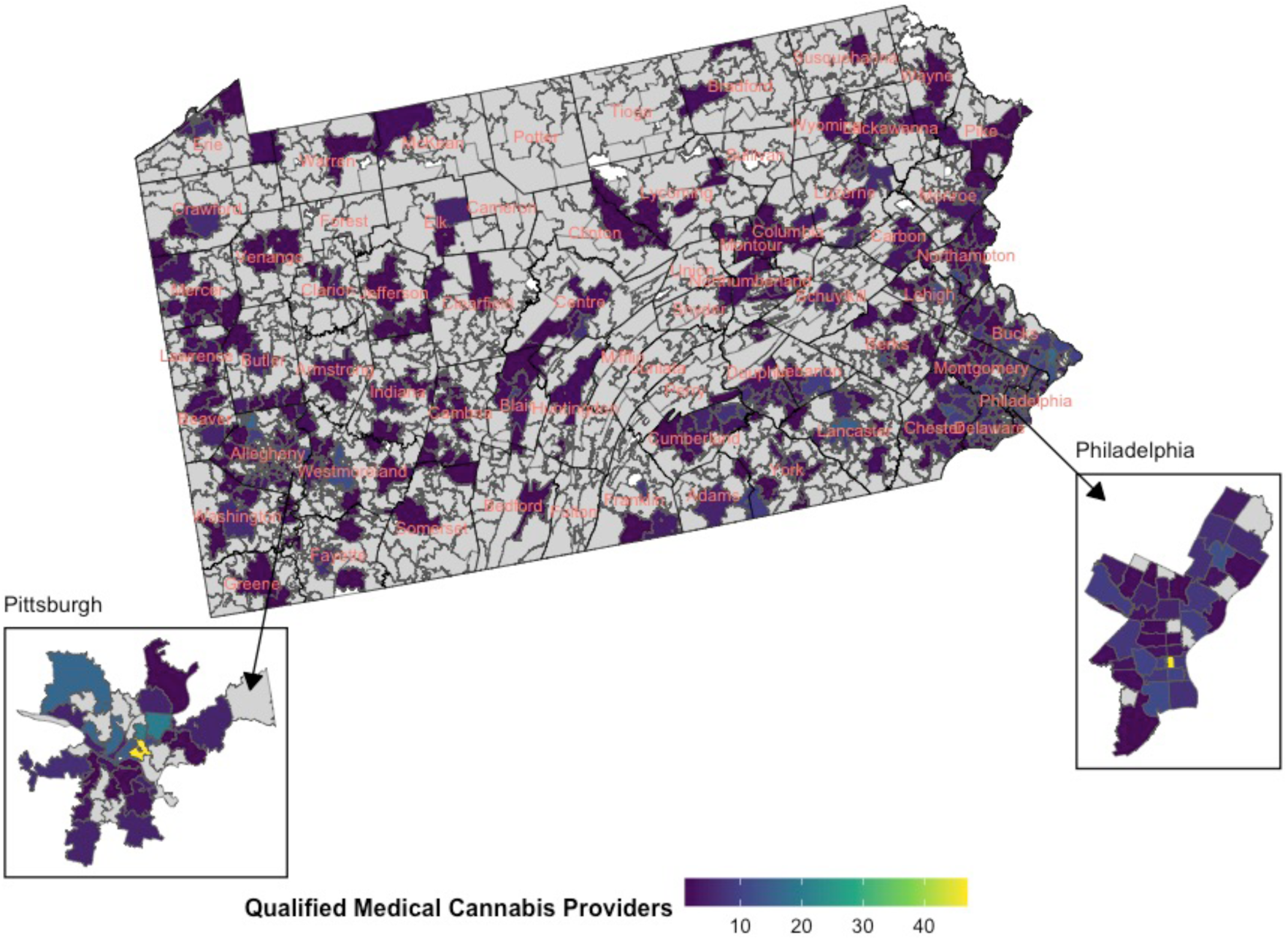
Distribution of qualified medical providers within Pennsylvania in 2022.Light gray indicates no patients were present in the zip code. White indicates limited data on population.

**Figure 4:**
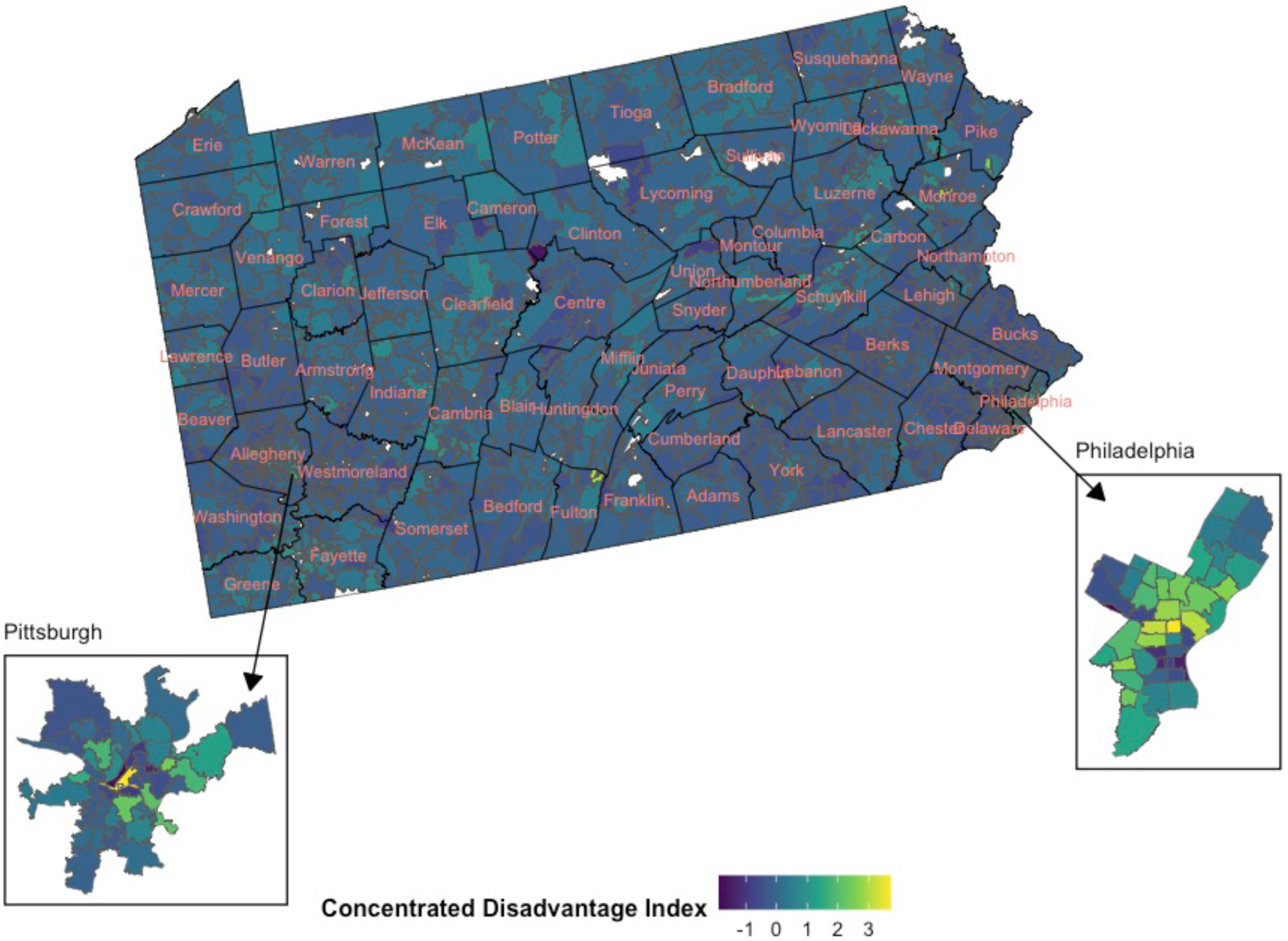
Distribution of concentrated disadvantage index within Pennsylvania in 2022. Light gray indicates no patients were present in the zip code. White indicates limited data on population.

Figure 3 illustrates the CDI at the zip code-level. The map reveals that areas with higher CDI scores are located across Pennsylvania, with some concentrations in urban centers like Philadelphia and Pittsburgh. The presence of high CDI values in both urban and rural areas indicates pockets of socioeconomic disadvantage across diverse geographic regions. Figure 4 shows the distribution of qualified in-clinic medical providers across zip codes. This figure highlights that the availability of qualified providers is unevenly distributed across the state, with significant concentrations in urban areas, particularly around Philadelphia and Pittsburgh. Rural areas and smaller towns show fewer providers, which might limit access to in-office services in those regions.

Table 1 presents the results of the Moran’s I Statistics, including Moran’s I, expectation value, variance, and p-value for both primary outcomes. Results suggest that the geographic distribution of both telemedicine-approved and all other patients is not random but rather exhibits a significant pattern of spatial clustering.

**Table 1:** Moran’s I Statistic for Primary Outcomes: Telemedicine Approved Patients and All Other Patients.

| Zip Code Level Variables | Moran's I | Expectation | Variance | p-value |
| --- | --- | --- | --- | --- |
| Telemedicine Approved Patients | 0.3488 | -0.0007 | 0.0003136 | <0.0001 |
| All Other Patients | 0.3509 | -0.0007 | 0.0003152 | <0.0001 |

**Note:** Moran's I statistics calculated at the zip code level. All p-values are significant at the $p < 0.0001$ level.

In Table 2, we see that the CDI does not have a statistically significant association with the incidence rate of telemedicine-approved patients. Without spatial lags, the IRR for the CDI is 0.988 (95% CI: 0.881, 1.107; *p* = 0.831), indicating no meaningful effect. When spatial lags are included, the IRR is slightly lower at 0.962 (95% CI: 0.885, 1.045; *p* = 0.355), but still not significant. The number of providers does not significantly affect the number of telemedicine-approved patients in either model. The IRR is close to 1 in both cases (IRR = 1.005, *p* = 0.344 without spatial lags; IRR = 1.007, *p* = 0.129 with spatial lags). The inclusion of spatial lags in the model is significant, with an IRR of 1.040 (95% CI: 1.037, 1.043; *p* =*<* 0.001).

**Table 2:** Associations between total telemedicine-approved medical cannabis patients and concentrated disadvantage index controlling for qualified medical cannabis providers, percent population white non-Hispanic, and spatial distribution.

| Zip Code Level Variables | Without Spatial Lags |  |  | With Spatial Lags |  |  |
| --- | --- | --- | --- | --- | --- | --- |
|  | IRR | 95% CI | P-value | IRR | 95% CI | P-value |
| Concentrated Disadvantage Index | 0.988 | 0.881, 1.107 | 0.831 | 0.962 | 0.885, 1.045 | 0.355 |
| Provider Counts | 1.005 | 0.994, 1.016 | 0.344 | 1.007 | 0.998, 1.017 | 0.129 |
| Percent White, non-Hispanic | 1.194 | 0.918, 1.555 | 0.187 | 0.929 | 0.749, 1.152 | 0.503 |
| Intercept | 0.001 | 0.001, 0.001 | 0.000 | 0.001 | 0.001, 0.002 | <0.001 |
| Spatial Lags | — | — | — | 1.040 | 1.037, 1.043 | <0.001 |

**Note:** Models are negative binomial generalized linear models with cluster robust standard errors clustered at the zip code level with a population-level offset.

In Table 3, we see that the CDI shows a significant negative association with the incidence rate of all other MC patients in both models. Without spatial lags, the IRR is 0.891 (95% CI: 0.839, 0.947; *p* =*<* 0.001),indicating that higher CDI values are associated with fewer all other MC patients in a given zip code. This negative association persists when spatial lags are included, with an IRR of 0.904 (95% CI: 0.855, 0.957; *p* =*<* 0.001). Provider counts also show a significant positive association with the number of non-Leafwell patients in both models, in contrast to Table 1. The IRR is 1.009 without spatial lags (*p* = 0.017) and remains consistent with spatial lags included (IRR = 1.009, *p* = 0.010). The inclusion of spatial lags is significant in this model as well, with an IRR of 1.002 (95% CI: 1.002, 1.003; *p* =*<* 0.001).

**Table 3:**
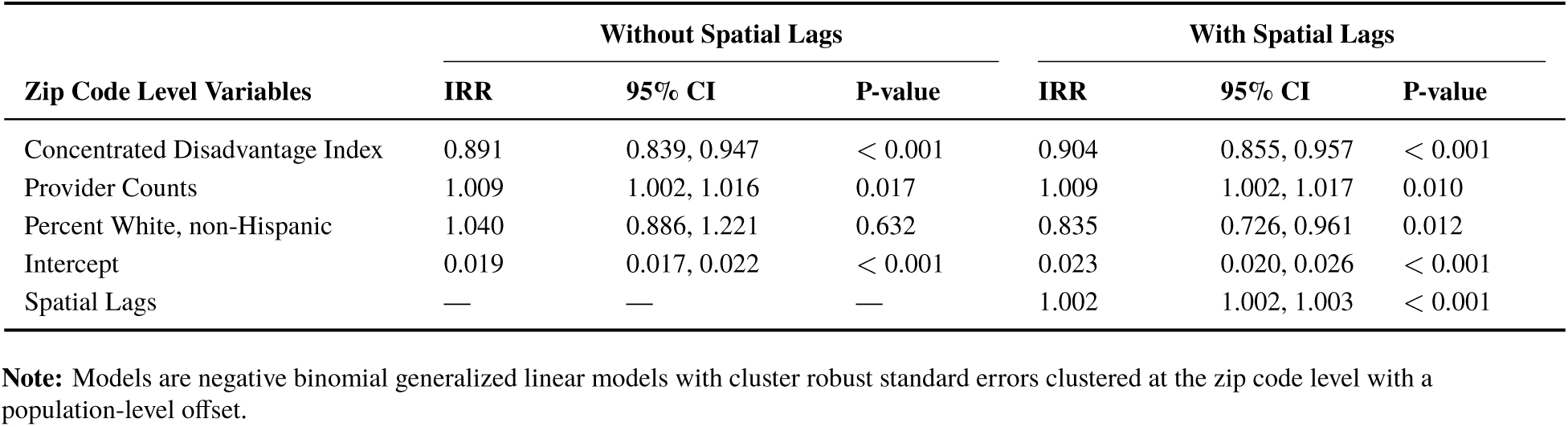
Associations between all other medical cannabis patients and concentrated disadvantage index controlling for qualified medical cannabis providers, percent population white non-Hispanic, and spatial distribution.

| Zip Code Level Variables | Without Spatial Lags |  |  | With Spatial Lags |  |  |
| --- | --- | --- | --- | --- | --- | --- |
|  | IRR | 95% CI | P-value | IRR | 95% CI | P-value |
| Concentrated Disadvantage Index | 0.891 | 0.839, 0.947 | < 0.001 | 0.904 | 0.855, 0.957 | < 0.001 |
| Provider Counts | 1.009 | 1.002, 1.016 | 0.017 | 1.009 | 1.002, 1.017 | 0.010 |
| Percent White, non-Hispanic | 1.040 | 0.886, 1.221 | 0.632 | 0.835 | 0.726, 0.961 | 0.012 |
| Intercept | 0.019 | 0.017, 0.022 | < 0.001 | 0.023 | 0.020, 0.026 | < 0.001 |
| Spatial Lags | — | — | — | 1.002 | 1.002, 1.003 | < 0.001 |

**Note:** Models are negative binomial generalized linear models with cluster robust standard errors clustered at the zip code level with a population-level offset.

## 4 Discussion

Using a combination of publicly available data from PA-DoH and proprietary data from Leafwell, we examined how the CDI, in-office MC provider counts, and racial composition influenced the distribution of our two outcomes, accounting for spatial autocorrelation. Our findings reveal significant differences in how socioeconomic factors and spatial distributions impact telemedicine-approved patients compared to all other patients. For telemedicine-approved patients, the CDI did not show a statistically significant association with the incidence rate of patients, indicating that telemedicine access mitigate some of the barriers typically associated with socioeconomic disadvantage. These findings suggest that telemedicine services, specifically for MC patients, may be more equitably distributed across socioeconomic strata than in-office MC providers alone, as found in Cunningham et al. (2022).

On the other hand, the CDI had a significant negative association with the incidence rate of all other patients, meaning that areas with higher socioeconomic disadvantage tended to have fewer all other MC patients. This contrast underscores a crucial finding: telemedicine may serve as a critical tool in reducing disparities around obtaining a qualification for a MC card, particularly in socioeconomically disadvantaged areas where traditional, in-clinic services are less prevalent or harder to access.

In the context of the current literature, our findings align with the Cunningham et al. (2022) study, which found disparities along socioeconomic lines in the distribution of in-office MC providers. Our research expands on this by differentiating between patients who obtained a medical card via a telemedicine provider and all other MC patients. Our findings, combined with Cunningham et al. (2022), suggest the benefits of MC may not be shared equitably across different zip codes.

As states attempt to address the inequities perpetuated by the war on drugs through legislative means, this knowledge can help policymakers better understand ways of alleviating the existing disparities around MC service access. A recent article found racial and ethnic differences in cannabis use following legalization among US states with MC laws. Martins et al. (2010) note that while white non-Hispanic and Hispanic individuals saw cannabis use increase, Black non-Hispanic individuals did not see similar changes. This difference may be the result of the persistent disparities in cannabis arrest rates (American Civil Liberties Union, 2020). However, part of this may be due to differences in accessing MC services found in Cunningham et al. (2022) and further discussed here. Our findings suggest that telemedicine services can alleviate some of the persistent disparities around MC access.

While our findings indicate that telemedicine can help alleviate some disparities in access to MC, the significant relationship between the CDI and the distribution of all other patients suggests that broader healthcare inequalities still persist within Pennsylvania. Policymakers in Pennsylvania, and policymakers in other states, should consider these disparities when crafting MC policies, ensuring that initiatives to expand telemedicine do not replace but rather complement efforts to improve access to in-person care, particularly in areas with high levels of concentrated disadvantage.

Telemedicine can address MC access challenges by connecting patients with cannabis-trained providers, regardless of location, thus improving access to expert advice in states with both medical and adult-use cannabis. It also helps bridge knowledge gaps among primary care providers by facilitating ongoing education and collaboration with specialists, ensuring cannabis use is safely integrated into treatment plans. Telemedicine enables better communication and continuity of care, keeping all healthcare providers informed and up-to-date on patient cannabis use, which reduces unrecorded or unauthorized prescription medication substitution. This approach increases patient confidence, supports personalized care, and enhances overall safety and efficacy in cannabis therapy.

### 4.1 Limitations

Several limitations must be considered when interpreting these findings. Geospatial analyses, while powerful, have inherent limitations related to the accuracy of spatial data and the potential for ecological fallacies. The use of zip code-level data, in particular, may mask important variations within smaller geographic areas, leading to oversimplified conclusions about the relationship between CDI and patient distribution. Additionally, the creation of the all other patients variable, which subtracts LPD patients from the total patient population, does not account for individuals who may have used both telemedicine and in-person services, nor does it account for patients who may have used another telemedicine service. This could potentially distort the findings, particularly if a significant number of patients utilized another telemedicine approval service to obtain their medical card. If this is true, the results may say more about telemedicine patients specific to the LPD rather than Pennsylvania as whole. Moreover, our study is cross-sectional, limiting our ability to infer causality from the observed associations. Longitudinal studies would be needed to fully understand how the relationship between CDI and patient distribution evolves over time, particularly as telemedicine continues to expand.

### 4.2 Conclusion

In conclusion, this study underscores the potential of telemedicine to mitigate healthcare access disparities in the context of MC. While traditional in-office MC providers appear to be less accessible in Pennsylvania, telemedicine shows promise in reaching populations that might otherwise be underserved.

## Data Availability

All publicly available data used as part of this study is available for use. Propriety data is not available.

## Notes

### Competing Interest Statement

This study was conducted using data provided by Leafwell, a Telehealth company that facilitates access to medical cannabis cards by connecting potential patients with qualified physicians. Leafwell does not manufacture or sell cannabis products. The authors declare no direct financial interests in the production or sale of cannabis products. However, as Leafwell provides services related to medical cannabis card certifications, the authors acknowledge that the findings of this study could indirectly benefit the company.

### Funding Statement

This study did not receive any funding

### Author Declarations

Ethics committee/IRB of Biomedical Research Alliance of New York (BRANY), IRB Number: IRB00000080, gave ethical approval for this work.

## References

American Civil Liberties Union (2020). A tale of two countries: Racially targeted arrests in the era of marijuana reform. Accessed: 2024-10-14.

AminiLari, M., Wang, L., Neumark, S., Adli, T., Couban, R. J., Giangregorio, L., Carney, C. E., and Busse, J. W. (2022). Medical cannabis and cannabinoids for impaired sleep: A systematic review and meta-analysis of randomized clinical trials. Sleep, 45(2):zsab234.

Boehnke, K. F., Sinclair, R., Gordon, F., Hosanagar, A., Roehler, D. R., Smith, T., and Hoots, B. (2024). Trends in u.s. medical cannabis registrations, authorizing clinicians, and reasons for use from 2020 to 2022. Annals of Internal Medicine, 177(4):458–466.

Bonn-Miller, M. O., Brunstetter, M., Simonian, A., Loflin, M. J. E., Vandrey, R., Babson, K. A., and Wortzel, H. (2022). The long-term, prospective, therapeutic impact of cannabis on post-traumatic stress disorder. Cannabis and Cannabinoid Research, 7(2):214–223.

Bradford, A. C. and Bradford, W. D. (2016). Medical marijuana laws reduce prescription medication use in medicare part d. Health Affairs, 35(7):1230–1236.

Bradford, A. C. and Bradford, W. D. (2017). Medical marijuana laws may be associated with a decline in the number of prescriptions for medicaid enrollees. Health Affairs, 36(5):945–951.

Bradford, A. C. and Bradford, W. D. (2018). The impact of medical cannabis legalization on prescription medication use and costs under medicare part d. The Journal of Law and Economics.

Bruce, D., Foster, E., and Shattell, M. (2021a). Perceived efficacy of medical cannabis in the treatment of co-occurring health-related quality of life symptoms. Behavioral Medicine, 47(2):170– 174.

Bruce, D., Grove, T. J., Foster, E., and Shattell, M. (2021b). Gender differences in medical cannabis use: Symptoms treated, physician support for use, and prescription medication discontinuation. Journal of Women’s Health, 30(6):857–863.

Cahill, S. P., Lunn, S. E., Diaz, P., and Page, J. E. (2021). Evaluation of patient reported safety and efficacy of cannabis from a survey of medical cannabis patients in canada. Frontiers in Public Health, 9:626853.

Chan, S., DeAngelis, C., Ganesh, V., Malek, L., Chow, E., O’Hearn, S., Blake, A., Wolt, A., Wan, B.-A., Zaki, P., Zhang, L., Lam, H., Slaven, M., and Shaw, E. (2017). Medical cannabis use for patients with post-traumatic stress disorder (ptsd). Journal of Pain Management, 10(4):385–396.

Charoenporn, V., Charernboon, T., and Mackie, C. J. (2023). Medical cannabis as a substitute for prescription agents: A systematic review and meta-analysis. Journal of Substance Use, 28(4):522–534.

Cunningham, C. O., Zhang, C., Hollins, M., Wang, M., Singh-Tan, S., and Joudrey, P. J. (2022). Availability of medical cannabis services by racial, social, and geographic characteristics of neighborhoods in new york: A cross-sectional study. BMC Public Health, 22(1):671.

DeWitt, S. (2020). Cannabis and coronavirus: Impact on medical cannabis industries in three states. SSRN Scholarly Paper ID 3689649.

Doucette, M. L., Casarett, D. J., Hemraj, D., Grelotti, D. J., Macfarlan, D. L., and Fisher, E. (2024a). Towards a comprehensive understanding of medical conditions among medical cannabis patients in a large database: A descriptive analysis. Population Medicine, 6:e71.

Doucette, M. L., Hemraj, D., Bruce, D., Fisher, E., and Macfarlan, D. L. (2024b). Medical cannabis patients under the age of 21 in the united states: Description of demographics and conditions from a large patient database, 2019–2023. Adolescent Health, Medicine and Therapeutics, 15:63–72.

Doucette, M. L., Macfarlan, D. L., Kasabuski, M., Chin, J., and Fisher, E. (2024c). Impact of medical cannabis treatment on healthcare utilization in patients with post-traumatic stress disorder: A retrospective cohort study. MedRXiv.

Drost, L., DeAngelis, C., Lam, H., Zaki, P., Malek, L., Chow, E., O’Hearn, S., Wan, B.-A., Blake, A., Chan, S., Wolt, A., Ganesh, V., Zhang, L., Slaven, M., and Shaw, E. (2017). Efficacy of different varieties of medical cannabis in relieving symptoms in post-traumatic stress disorder (ptsd) patients. Journal of Pain Management, 10(4):415–422.

Drug Policy Alliance (2021). Inequitable marijuana criminalization, covid-19 and socioeconomic disparities: The case for community reinvestment in new york. Accessed: 2024-10-16.

Ebbert, J. O., Ramar, P., Tulledge-Scheitel, S. M., Njeru, J. W., Rosedahl, J. K., Roellinger, D., and Philpot, L. M. (2023). Patient preferences for telehealth services in a large multispecialty practice. Journal of Telemedicine and Telecare, 29(4):298–303.

Erridge, S., Holvey, C., Coomber, R., Hoare, J., Khan, S., Platt, M. W., Rucker, J. J., Weatherall, M. W., Beri, S., and Sodergren, M. H. (2023). Clinical outcome data of children treated with cannabis-based medicinal products for treatment resistant epilepsy—analysis from the uk medical cannabis registry. Neuropediatrics, 54(3):174–181.

Erridge, S., Kerr-Gaffney, J., Holvey, C., Coomber, R., Barros, D. A. R., Bhoskar, U., Mwimba, G., Praveen, K., Symeon, C., Sachdeva-Mohan, S., Sodergren, M. H., and Rucker, J. J. (2022). Clinical outcome analysis of patients with autism spectrum disorder: Analysis from the uk medical cannabis registry. Therapeutic Advances in Psychopharmacology, 12:20451253221116240.

Fairman, B. J. (2016). Trends in registered medical marijuana participation across 13 us states and district of columbia. Drug and Alcohol Dependence, 159:72–79.

French, M. T., Zukerberg, J., Lewandowski, T. E., Piccolo, K. B., and Mortensen, K. (2022). Societal costs and outcomes of medical and recreational marijuana policies in the united states: A systematic review. Medical Care Research and Review, 79(6):743–771.

Gajarawala, S. N. and Pelkowski, J. N. (2021). Telehealth benefits and barriers. The Journal for Nurse Practitioners, 17(2):218–221.

Ghimire, K. M., . M. J. C. (2020). Medical marijuana and workers’ compensation claiming. Health Economics, 29(4):418–434.

Harris, M., Erridge, S., Ergisi, M., Nimalan, D., Kawka, M., Salazar, O., Ali, R., Loupasaki, K., Holvey, C., Coomber, R., Usmani, A., Sajad, M., Hoare, J., Rucker, J. J., Platt, M. W., and Sodergren, M. H. (2022). Uk medical cannabis registry: An analysis of clinical outcomes of medicinal cannabis therapy for chronic pain conditions. Expert Review of Clinical Pharmacology, 15(4):473–485.

Jing, F., Li, Z., Qiao, S., Zhang, J., Olatosi, B., and Li, X. (2022). Investigating the relationships between concentrated disadvantage, place connectivity, and covid-19 fatality in the united states over time. BMC Public Health, 22(1):2346.

Leafwell (2024). Medical cannabis research dashboard. https://leafwell.com/research/dashboard. Accessed November 10, 2024.

Mahabir, V. K., Merchant, J. J., Smith, C., and Garibaldi, A. (2020). Medical cannabis use in the united states: A retrospective database study. Journal of Cannabis Research, 2:32.

Mahon, E. (2023a). Get the data: Spotlight pa makes info on why patients qualify for medical marijuana publicly available. Spotlight PA.

Mahon, E. (2023b). Spotlight-pa-marijuana-certifications [computer software]. Spotlight PA. Original work published 2023.

Mahtta, D., Daher, M., Lee, M. T., Sayani, S., Shishehbor, M., and Virani, S. S. (2021). Promise and perils of telehealth in the current era. Current Cardiology Reports, 23(9):115.

Mangoo, S., Erridge, S., Holvey, C., Coomber, R., Barros, D. A. R., Bhoskar, U., Mwimba, G., Praveen, K., Symeon, C., Sachdeva-Mohan, S., Rucker, J. J., and Sodergren, M. H. (2022). Assessment of clinical outcomes of medicinal cannabis therapy for depression: Analysis from the uk medical cannabis registry. Expert Review of Neurotherapeutics, 22(11–12):995–1008.

Manson, S., Schroeder, J., Van Riper, D., Knowles, K., Kugler, T., Roberts, F., and Ruggles, S. (2023). Ipums national historical geographic information system: Version 18.0 [dataset]. IPUMS.

Marcondes, F. O., Normand, S.-L. T., Le Cook, B., Huskamp, H. A., Rodriguez, J. A., Barnett, M. L., Uscher-Pines, L., Busch, A. B., and Mehrotra, A. (2024). Racial and ethnic differences in telemedicine use. JAMA Health Forum, 5(3):e240131.

Martins, S. S., Ghandour, L. A., Lee, G. P., and Storr, C. L. (2010). Sociodemographic and substance use correlates of gambling behavior in the canadian general population. Journal of Addictive Diseases, 29(3):338–351.

National Academies of Sciences (2017). The Health Effects of Cannabis and Cannabinoids: The Current State of Evidence and Recommendations for Research. National Academies Press.

National Conference of State Legislatures (2024). State medical cannabis laws. NCSL.

New Jersey Cannabis Regulatory Commission (2024). Cannabis related laws. NJCRC. Accessed: 2024-10-14.

Nicholas, M., Erridge, S., Bapir, L., Pillai, M., Dalavaye, N., Holvey, C., Coomber, R., Rucker, J. J., Weatherall, M. W., and Sodergren, M. H. (2023). Uk medical cannabis registry: Assessment of clinical outcomes in patients with headache disorders. Expert Review of Neurotherapeutics, 23(1):85–96.

Omboni, S., McManus, R. J., Bosworth, H. B., Chappell, L. C., Green, B. B., Kario, K., Logan, A. G., Magid, D. J., Mckinstry, B., Margolis, K. L., Parati, G., and Wakefield, B. J. (2020). Evidence and recommendations on the use of telemedicine for the management of arterial hypertension. Hypertension, 76(5):1368–1383.

O’Connell, M., Sandgren, M., Frantzen, L., Bower, E., and Erickson, B. (2019). Medical cannabis: Effects on opioid and benzodiazepine requirements for pain control. Annals of Pharmacotherapy, 53(11):1081–1086.

Pankratz, T. (2023). Impact of Telehealth Versus Traditional Visits on Pain Management Patients Utilizing Medical Cannabis During the COVID-19 Pandemic. PhD thesis, Walden University.

Pennsylvania Department of Health (2024). Department of health medical marijuana approved practitioners.

Pillai, M., Erridge, S., Bapir, L., Nicholas, M., Dalavaye, N., Holvey, C., Coomber, R., Barros, D., Bhoskar, U., Mwimba, G., Praveen, K., Symeon, C., Sachdeva-Mohan, S., Rucker, J. J., and Sodergren, M. H. (2022). Assessment of clinical outcomes in patients with post-traumatic stress disorder: Analysis from the uk medical cannabis registry. Expert Review of Neurotherapeutics, 22(11–12):1009–1018.

Purcell, C., Davis, A., Moolman, N., and Taylor, S. M. (2019). Reduction of benzodiazepine use in patients prescribed medical cannabis. Cannabis and Cannabinoid Research, 4(3):214–218.

R Core Team (2024). R: A language and environment for statistical computing. R Foundation for Statistical Computing.

Shaver, J. (2022). The state of telehealth before and after the covid-19 pandemic. Primary Care, 49(4):517–530.

State of Connecticut (2024). Policies and procedures for connecticut’s adult-use cannabis program. CT.Gov. Accessed: 2024–10-14.

Tchero, H., Kangambega, P., Briatte, C., Brunet-Houdard, S., Retali, G.-R., and Rusch, E. (2019). Clinical effectiveness of telemedicine in diabetes mellitus: A meta-analysis of 42 randomized controlled trials. Telemedicine Journal and E-Health, 25(7):569–583.

Ullman, D. L. (2017). The effect of medical marijuana on sickness absence. Health Economics, 26(10):1322–1327.

Venables, W. N. and Ripley, B. D. (2002). Modern Applied Statistics with S. Springer, 4th edition.

WHYY (2024). Pa. marijuana laws: Here’s what you need to know. WHYY. Accessed: 2024-10- 14.

